# A comprehensive estimation and analysis of the basic reproduction number (R0) of novel corona virus in India: A comparative study with different lockdown phase of COVID-19

**DOI:** 10.1101/2020.07.10.20150631

**Authors:** Tushant Agrawal, Mayank Chhabra

## Abstract

**Background:** World Health organization declared Covid-19 as an outbreak, hence preventive measure like lockdown should be taken to control the spread of infection. This study offers an exhaustive analysis of the reproductive number (R0) in India with major intervention for COVID-19 outbreaks and analysed the lockdown effects on the Covid-19.

**Methodology:** Covid-19 data extracted from Ministry of Health and Family Welfare, Government of India. Then, a novel method implemented in the incidence and Optimum function in desolve package to the data of cumulative daily new confirmed cases for robustly estimating the reproduction number in the R software.

**Result:** Analysis has been seen that the lockdown was really quite as effective, India has already shown a major steady decline. The growth rate has fluctuated about 20 percent with trend line projections in various lockdown. A comparative analysis gives an idea of decline in value of R0 from 1.73 to 1.08. Annotation plot showing the predicted R0 values based on previous lockdown in month of June and July.

**Conclusion:** Without lockdown, the growth might not have been contained in India and may have gone into the exponential zone. We show that, the lockdown in India was fairly successful. The effect partial lifting of the lockdown (unlock) is also seen in the results, in terms of increment in R0 values. Hence this study provides a platform for policy makers and government authorities for implementing the strategies to prevent the spread of infection.

## Introduction

Corona virus disease COVID-19 seems to be more infectious than the previously identified SARS viruses and is caused by a member of the viral SARS family, named the Corona virusCoV 2 virus. COVID is linked to a human respiratory disorder, which in the first quarter of 2020 has been declared by the World Health Organization as a global pandemic [1]. Specific policy measures, such as wearing of masks and using face shields [2, 3] contained social distancing, responded to this extraordinary situation by the global community. Governments around the world made more severe and perhaps onerous lockdown steps [4] as a policy measure of containment [5].The number of individuals who are susceptible to infection is the average number represent as reproduction number. This means that the number of healthy individuals afflicted by the total count of infectedis reproduced. The basic reproductive number, R0, specifies the intensity of transmissible infectious diseases and thus provides basic knowledge essential for health care interventions. The basic reproduction R0 is the number of secondary infections by a disease’s primary infection, and it is a basic epidemiological metric that assesses the first-rate of infection transmission throughout a disease outbreak, along with other aspects [6]. The number of communications, in particular, and the effective number [7] of vulnerable interactions per possible infectious are greatly decreased by various policy enforcing and steps of mitigation. R0 predictions are based upon the assumption that perhaps the values at the beginning of the outbreak represent the population’s assets and thus predict a plausible rate of infectious disease if an outbreak is back on the rise. The infection occurs to start growing in a community whenever R0>1, and not after R0<1. The bigger the significance of R0, the more difficult the outbreak is to manage and the more likely an epidemic is to occur.

More than half million people are affected infected with the Novel Corona virus in India according to the latest figures from MoHFW of India [8] and other tracking websites [9]. Several medical professionals across the country proposed the idea of social distance since it was necessary for the intimate contact link between humans to be disrupted. Announcing regional lockdowns was the very first step in the right direction. With regard to cases in Fig. 1, it is indeed obvious that a spike was noted on 21 March 2020 in India. Thereafter, on 22 March 2020, the government formally declared a freeze, planning the situation in the manner of the government, and in Lockdown, people were going to be managed. It clearly demonstrates that Indian government were responsive enough even to estimate the spread rate in the India and to take the required measures to maintain social distance by declaring a restrictive locking phase. The Indian government has taken this initiative strictly and announced the 25 March 2020 lockdown, which had lasted 21 days until 14 April 2020.

**Fig 1.**
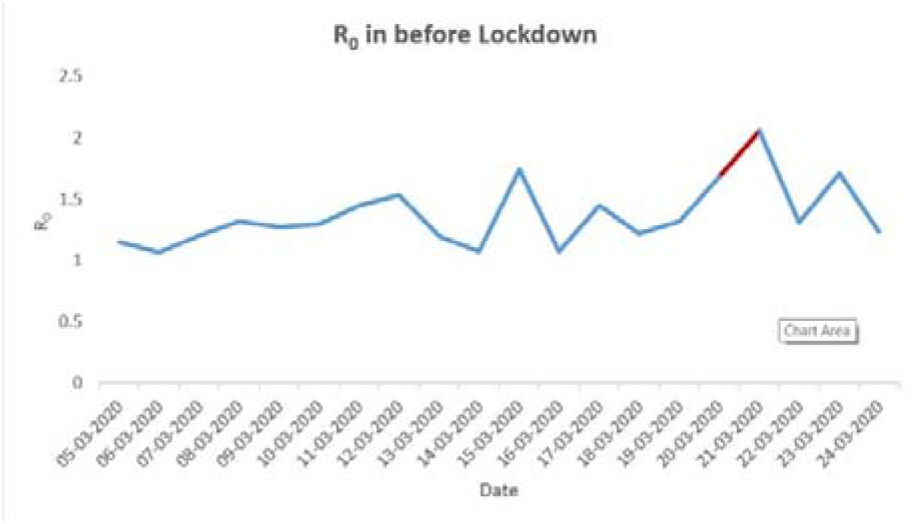
Estimated R0 values from 5 March to 24 March (pre-lockdown)

The residents were only permitted to leave in emergencies with prior authorization from the local authorities. In order to straighten the trajectory of the afflicted case and restrain the rapid growth of patients in India, all these instructions had been given. The first locking had been further prolonged with a short period from 15 April 2020 until 3 May 2020, 3 May 2020 until 17 May 2020, and from 18 May to 31 May 2020, on the report, based on the first lockdown and showing COVID-19 scenario. India’s lock-down period has been disrupted in recent days by two big incidents that have linked employees and staff from one region to another in Delhi, leading to a growing number of cases in different regions of India. At this time, it was through promotional activities and numerous participation initiatives that the prime minister of India tried to bind the Indian people across its entire country. It is crucial that we research the present predicament and consequences of different such events in India by means of data analysis based on the mathematical parameters in India at this moment. This research aims to examine the propagation of COVID-19 in India at a systematic level and the effects of different policies that are implemented by the government.

This study offers an exhaustive analysis of the reproductive number (R0) in India with major intervention for COVID-19 outbreaks and analysed the lockdown effects on the Covid-19 viral disease spread. The cases are growing rapidly and the Indian administration department must implement combative preventive measures. In this study, various facets are addressed, including the spread of diseases in various lockdown in India, predicted reproductive numbers for the next several days, the effect of this forecasting on prolonged lockdown, the influence of social distances on the residents in India, and the analysis of Indian lock-up methodologies. It has also studied the potential justification for such defections. Certain mathematical equations and parameters for the number reproduction (R0) have been implemented in the current analysis.

## Review of Literature

A three-week nation-wide lockdown had been implemented on 25th March 2020 to reduce the propagation of the Coronavirus. This is a tough decision for any nation, especially for a country with 22 percent of the population living below the poverty line and 90 percent of its workforce working in the informal sector. In the informal economy, approximately 400 million laborers are in danger of extreme poverty during the catastrophe (11). For one in six urban residents, living standards are comparatively despicable and the density of slum areas is a major problem and obstacle for keeping people socially distant. The aim of the national lockdown is to prevent the spread of Coronavirus so that the government can adopt a multi-pronged policy: append more beds towards its number of hospitals, level up the development of COVID-19 testing kits and personal protection equipment (PPE) for healthcare workers. This lockdown tends to help break the pattern of infections. It expects to defer the disease progression until the health infrastructure is able to cope with the sharp rise in cases. Quick screening at the start of transmission has been one of the key factors responsible for dropping Rt, as it attempts to determine and isolate patients at an initial point. That being said, India has limited facilities, which means that adequate assessments could not be carried out in some regions; as a consequence, they have done little to prevent the transmission of the infection. In addition to the national lock-up, social-welfare programs have been implemented although not in an uniform manner by all regions. Policy making is an important aspect at the state level to monitor the transmission of infection, as it reassures us how effective the government is in attempting to implement and impacting laws and regulations. Healthcare services is also a crucial part of building the regulations on health care delivery and social assistance processes. It is formulated that such attributes get a positive bond to the downward trend of Rt. Initially, to acknowledge how appropriate action are being enforced to monitor the epidemic, and furthermore, to provide us with essential information about whether the government could perhaps raise or minimize prohibitions on the basis of contending economic growth and human security objectives. It is anticipated that a nationwide lockdown would have been crucial in enhancing down the value of Rt.

Singh and Adhikari, have used R0 of 2.10, revealed that the approximated infective in India is probable to be 900 million in the lack of mention of any social interactions or closure (12). Schueller et al. study estimated that, with pretty tough lockdown and increased social distancing, India will have had a peak sum of 97 million infections and that the number of infections is inclined to be more than 1100 million by September (13). Dwivedi et al., using a mathematical estimation, projected an R0 of 1.56 for the period 4-19 April 2020 (14). Thus according Singh and Adhikari (2020), the estimated R0 varied between 2.10 in March 2020 to 0.50 in August 2020 (2). Sangeeta Bhatia et al. argues that short-term estimates by Imperial College London recommend a weekly rate of 1.45 for India beginning on May 03, 2020 (15). Kissler et al. have said that the optimum winter time for R0 is 2.0 and for summer time it is 1.4 (16). Statistics from Wuhan China propose that average daily reproduction decreased from 2.35 to 1.05 after travel restrictions, home quarantine, centrally controlled quarantine and medication, and improved healthcare resources (17-18). The projected R0 for the United States of America had been 1.51 on 1 April 2020 (19). In the United States, the time-varying reproduction number decreased from 4.02 to 1.51 between 17 March and 1 April 2020 (9). Italy registered a reproduction number of 2.6 and 3.3, assuming that the transmission dates from 5 February 2020 and 10 February 2020 respectively (20). The number of reproductions in the Republic of Korea has been projected at 2.6 and 3.2 (as of 5 March 2020) with an assumed transmission starting on 31 January 2020 and 5 February 2020 (10) respectively. Literature suggests the mean serial interval for COVID-19 ranges from 4 to 8 days (21-24). Recent analyses by Duet al. used a much larger sample that includes up to 468 pairs, making their estimates of between 4 and 5 days which are more statistically reliable (21). The estimated mean serial interval is shorter than the preliminary estimates of the mean incubation period (approximately 5 days) (23-24). When the serial interval is shorter than the incubation period for infectious disease, the pre-symptomatic transmission is likely to have taken place and may occur even more frequently than symptomatic transmission (25). Results demonstrate that all stages of lockdown have led to a decrease in the reproduction number of India, which really is similar to the findings of such studies (14, 26). Even so, such strategies could not actually work, as India is a multicultural nation in which some countries are highly sophisticated in terms of health infrastructure and human improvement, whereas some countries are stagnating behind in these amenities. According to the Goli et al study, India is only detecting 3.6 per cent of the total number of COVID-19 infections throughout its regions (27). They often suggest that India needs to boost its inspection capabilities and carry out pervasive testing, as late detection of the virus brings patients in bigger need of ventilation and ICU care, that inflicts increased expenses on health services. India also should take randomized population-level tests to evaluate the pervasiveness of disease. In applied to measure the exact prevalence of COVID-19 infections in India, the well-established National Family Health Survey (NFHS) structure can be adopted as an analysis to reach the prevalence and severity of COVID-19 (28). Six objectives of the research are broadly defined in the research studies by Rajan et al. (29). It has already been noticed that rate of growth of infected cases has been monitored with the support of National Lockdown, even though some unrestrained mass level incidents might have adversely affected the infected cases. Tactics for the prolongation of the lockdown have been reviewed and summarized. It did appear that certain vital services ought to be available to the Indian citizens, and the national lockdown should take place over the next 2-4 weeks. In addition, they even said that without the need for a lockdown, increase may have not been usually contain in India and it may have entered the exponential zone too rapidly.

Fortunately, maybe there are other aspects, such as the comeback of migrant workers to their homes, effective contact tracing and reliability of quarantine centres, which lead to the pathogenesis of infection but it couldn’t be assessed in the study. In the upcoming days, confirmed cases are projected to increase daily in India as migrants move back home in huge numbers. The number of insulation beds, ICU beds and ventilator beds should really be nearly doubled in all regions. This requires to be improved in order to facilitate a large number of cases in the upcoming days, whenever the daily confirmed cases will be at their peak, as the health infrastructure is not up to the standard, even in developed nations, to track with the outbreak.

## Methodology

The global pandemic reproduction number (R0) can be defined in various ways, for example with the Stochastic SIR (DCM) model, the Poissonian probability-based (ML) model, and the Exponential Growth Rate (EGR) model. In the analysis of reproduction numbers with a total daily basis case, we have used the Stochastic dynamic contact model-based method [10].

By contrast to the traditional linear system, a Susceptible-Infected-Recovered (SIR) model is taken into account in that it adapts to increased uncertainty and better quantifies uncertainties of this number relative to the traditional linear system. In this context, the population susceptible, infection and recover are indicated by S(t), I(t) & R(t), and that the overall population of the demographic is the number of N = S(t)+I(t)+R(t).

The infectious period of an infected individual is a random variable T ∼ exp(γ) & the reproduction rate is 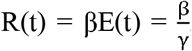, where β, the infection rate that manage the transition between S and I, and γ, the removal or recovery rate that manage the transition between I and R. The mathematical essence of the model can captured by a set of coupled first order linear homogenous differential equations given such to be:

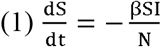

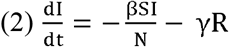

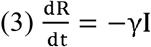

With above equation calculated R0 value, we implemented some mathematical algorithm and method (LBFGS) with packages of R programming language code and demonstrate the optimal values of β and γ parameters for daily basis cases. The ode() function that defined ordinary differential equation, from r packages deSolve creates equation easy for the system and what we want to estimates values of parameters for calculation of R0 for the current study, simply used the optim() function built into R base. We can compute it in R by given below code:

Opt_par

R0<-as.numeric(Opt_par[1] / Opt_par[2])

R0

The Limited-memory Broyden-Fletcher-Goldfarb-Shanno (L-BFGS) [30] algorithm has been used in scenarios of outbreak where both the objective function and its gradient are analytically measured for resolving high-dimensional minimization issues. The L-BFGS algorithm is a class of quasi-Newton optimization procedures that fix the minimum problems in the Hessian matrix of the optimal solution.

The value of reproduction number is lower (1.00<R0<2.5) indicate a fact that the number of infected cases stayed constant that would be equal to 1 in the beginning of the outbreak. In current study, we can state that reproduction number (R0) 1.7 mean on average 1.7 individual could infected through single infected individual. Through a formula [1-(1/R0)], which indicate the population would be immunized to stop the transmission of the ailments. However, R0 is 1.4 that should be show that 28% of the community have been immunized to stop the transmission of the outbreak of COVID-19.

## Result

The nationwide lockdown on 22nd March 2020 announced by the central government of India, proves to be an effective measure in reducing the value of reproduction number (R0) (Fig 1). Merely after this effective measure, a lockdown for 21 days was announced restricting almost all services (except emergency and essential services) to break off the spread of covid-19 infection and in hope to flattened the curve of infection. Initially at the beginning of lockdown i.e. on 25 March, the R0 value is around 1.37, thus merely started declining within 2 days of lockdown. During first week of April around two sharp peaks were obtained indicating the increase in number of infected cases, this is due to Tablighi Jamaat religious congregation that took place in Delhi during March 2020. More than 9,000 people have attended the congregation, from various states of India, and with more than 4,000 confirmed infected cases [31] and around 27 deaths linked to this event were reported. By 3 April, over 950 confirmed cases were detected across 14 states and union territories in the country, including 97 percent of the total confirmed cases in the country (647 out of 664 cases) [32]. Tamil Nadu was the worst affected state, as 364 of the 411 people who tested positive had attended the event, 259 of the 386 cases in Delhi and 140 of the 161 cases in Andhra Pradesh were linked to the event [33]. According to health authorities, until 2 April, among 2000 positive cases in India nearly 400 cases can be epidemiologically traced to the Tablighi Jamaat cluster [34]. As of 4 April, 1,023 cases with links to this cluster were reported which is about 30% of total cases in the country [35].

Afterward the value of R0 peak declined continuously to 1.21, hence reducing the spread (Fig 2). Henceforth lockdown 1 can be considered as significantly effective as it not only control the spread of infection but also reduces the reproduction number from 1.37 to 1.23. Hence without proper lockdown measures, the spread rate may not contained in India and might have gone into the exponential growth rapidly. This gives all the state level and national level authorities and health workers to get prepared for the rising number of cases.

**Fig 2.**
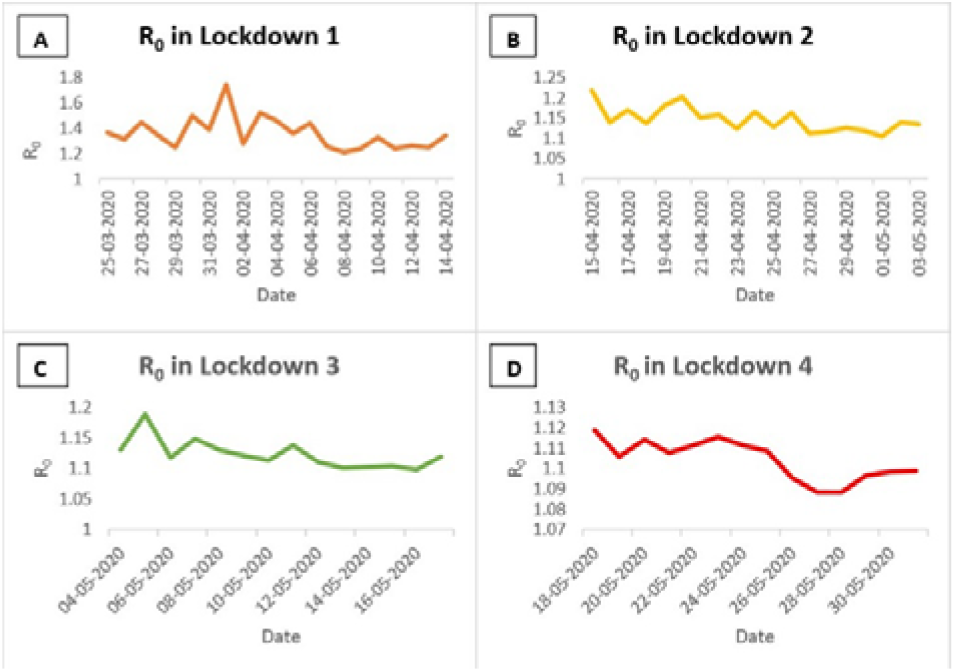
Plot showing the Scenarios of Different lockdown with R0 values of COVID-19 in India

Effectiveness of lockdown 1 gives a decision to healthcare authorities and central government to implement lockdown 2 to further reduce the spread of infection. Lockdown 2 was implemented for 18 day from 15^th^ April until 3^rd^ May with similar restricted movement as in lockdown 1. Around 18 April a peak was observed where R0 value increases due to the fall off effect of tablighi jamaat, Union Health Ministry said 4,291 out of 14,378 confirmed cases in 23 Indian states and union territories have been linked to this event, around a third of all cases [8]. Afterwards the R0 peak continuously declined from 1.21 to 1.13, hence this can be considered as an effective lockdown in contrast to preventing the spread of infection. Both lockdown 1 and lockdown 2 last for almost 39 days with restricted movement, this causes an uncertainty and lack of household resources, tends public and authorities to generate an alternative options. During lockdown 3 the state government allow a flow of movement in orange and green zone areas for a restricted time and with complete precaution measures like social distancing, wearing mask etc. Moreover the transportation restrictions are somehow lifted with shramik train facilities and pass facility for personal vehicle; this leads to a sudden rise in peak around 5^th^ May, which drastically goes down as the precautionary measures are taken carefully. Furthermore, lockdown 3 doesn’tprove as an effective because the restricted policies are lifted and a slight change in spread rate is observed i.e. from 1.13 to 1.12 value of R0.

Unlike lockdown 3, a considerably change in spread of infection was observed further during lockdown 4 i.e. from 18 May till 30^th^ May, the R0 value decreased down from 1.12 to 1.08 (Fig 2). This is due to health belief of people due covid-19 like wearing mask, good hygiene practice, social distancing and other factors. Hence a comparative analysis of all lockdown based on R0 value was shown in Fig 3.

**Fig 3.**
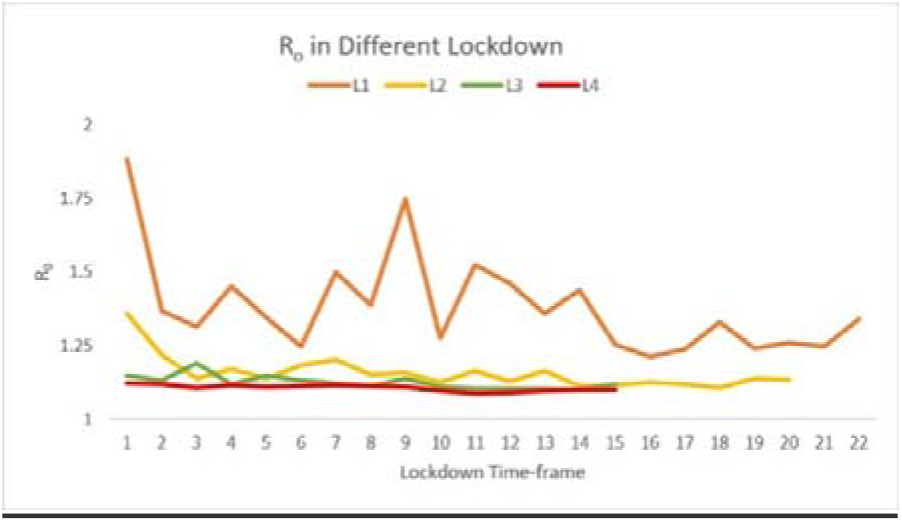
A comprehensive analysis chart define the transmission of corona virus reproduction rate (R0) in varies lockdown time frame in India

Taking lockdown into account, it gives a continuously decline in R0 value from 1.73 to 1.08 from lockdown 1 to lockdown 4. Hence using a predictive analytics, a prediction by regression modelling can give us a brief idea about the decline in spread rate (R0) in next consecutive lockdown in month of June i.e. lockdown 5. It was observed a sudden decline in R0 value to less than 1 in the end of June if lockdown 5 was implemented (Fig 4). The R0 value less than 1 simplifies the spread rate is optimal and the infection curve will flattened, with low spread rate. This prediction analysis suggested that lockdown in June month could defiantly decline the number of cases and leads to fall the peak, forming a bell shaped curve.

**Fig 4.**
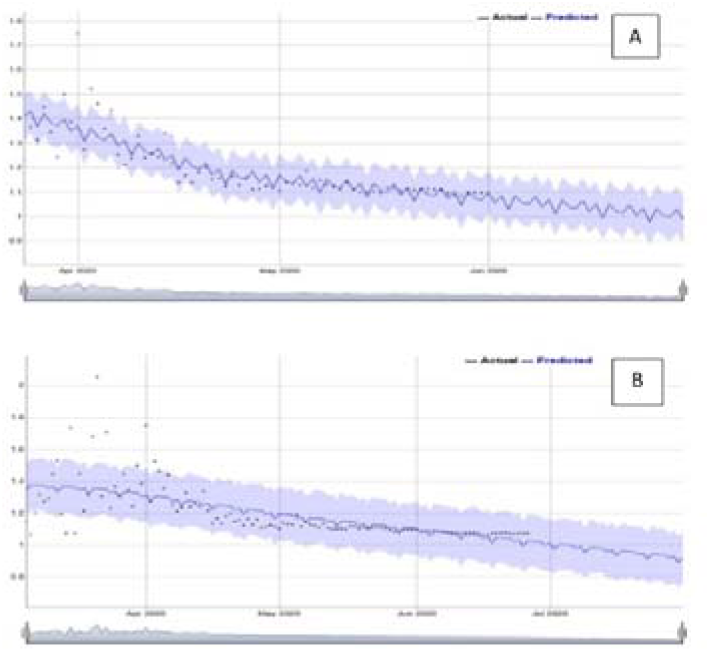
Annotation plot showing the predicted R0 values based on calculated R0 values by daily cumulative cases A) Prediction of R0 values for month of June (if lockdown sustain) based on calculated R0 values from 24 march till 31 May and B) Prediction of R0 values for month of July (if lockdown sustain) based on calculated R0 values from 5 march 2020 till 26 June.

**Fig 5.**
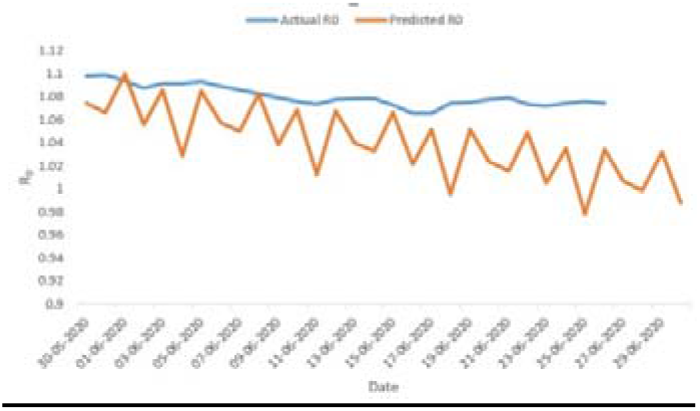
Comparison between actual (without lockdown) and predicted R0 values (if lockdown sustain in month of June) for month of June.

## Discussion

It has been seen that the lockdown was really quite as effective as a quarantine criterion. From 25 March 2020, the lockdown in India began.Within few days of the commencement, in the instant reproductive number, India has already shown a major steady decline.The growth rate has been calculated as the difference in number of cases between two consecutive days divided by the count of infected cases on the previous day of the two days and the reproduction number (R0) has been defined with daily basis cumulative cases thatare put in the computational algorithm and on the basis of infection rate and therecovery rate parameter has been calculated the value of R0. Although the number had been in single digits in the early stages, the growth rate was very high. Neglecting this duration, the growth rate has fluctuated about 20 percent with a trend line projections of up to about 28 percent, taking into account the second stage of the phase from 5 March to 22 March 2020, precisely well before lockdown. The growth rate, meanwhile, marginally increased for the duration after lockdown, i.e. from 22 March 2020, but still about the same 20 percent level. And the trend line also expected a moderate growth of 28 percent per day [36]. Furthermore, the next lockdowns shows only slight changes in terms of declining R0 value.

Fu-Chang Hu et al suggested that the lockdown of Wuhan on January 23, 2020, was a very smart, brave, and quick move by computing R0(t) values, hence the estimated R0(t) began to drop just 2−4 days after the lockdown and the trend of the computing R0(t) was declining monotonically for about 10 days. Two weeks later, the estimated R0(t) surprisingly reduced to the level below 1.0 around February 17−18, 2020, In the end, China would win the battle against the corona virus as long as its resurgence, if any, is well managed. We believe that it is much more important to see the approximate R0(t) decline than to see the decrease in the regular number of newly reported cases during an ongoing infectious outbreak. And, the consistent decline of the estimated R0(t) in trend over time is more important than the actual values of the estimated R0(t) themselves. The steeper the slope, the sooner the epidemic ends [37].

Even so, recent results have been consistent with previous COVID-19outbreak predictions in India of R0.The only tactic to maintainthe epidemic effectively is the non-pharmaceutical strategies. The first criterion is that theclinics should be equipped to deal with patients with minimal pain, speedy recovery andlowest mortality. The second most significant requirement is to plan community for theproliferation of contamination by providing insulation stations, quarantine centres, and community distances [38]. Recognizing contaminated cases is the key to certain mitigation but it is amajor drawback that standard diagnostic kits are not available and are not affordable.Annual meantemperature and successful social distance in one nation would probably have a substantialsuppressive influence on COVID-19 propagation. High temperatures and moisture have beenindicated in a study conducted to decreases the daily new cases and new death [39]. In India, theCOVID-19 epidemic has been monitored with the government actions on demographiccontrol, isolation of verified infectious persons, quarantine of suspected infected personsbased on touch monitoring, hospital and health care, social separation initiatives, communitysocio-economic structure, lock-down, and policies, might well predict the establishment ofnew cases, recoveries and death, etc.

India is currently minimized lockdown restrictions and choosing partial lockdowns as a tacticfor phasing out the lockdowns. In certain cases, largely due to ignorance ormisunderstanding, the infected persons & belongings remain ignored and are found by theGovernment after a surveillance campaign. Unanticipated labour immigration from cities torural ancestral lands can be shifting the area of infection in most scenarios. Anyone of theseactivities tends to affect the differential population distribution of infected and dead cases.

## Conclusion

The whole study provided an overview of the COVID-19 lockdown measure in India. The increase in the number of cases of COVID-19 could be contained under national lockdown. The development could not have been controlled in India without the need for lockdown and may have been too rapidly explosive. This helps both state-level and regional managers and health professionals to brace themselves for the growing number of cases.We have shown that in India the lockdown has been very effective without Tablighi jamaat, migration, or enhanced transmission in some areas, etc. Moreover predictive analysis for further implementation of lockdown was also shown with R0 values for moth of July. The effect partial lifting of the lockdown (unlock) is also seen in the results, in terms of increment in R0 values. Hence this study provides a platform for policy makers and government authorities for implementing the strategies to clog the transmission of infection.

## Data Availability

Covid data is taken from Ministry of health and family welfare (MoHFW) website and processed using R software desolve package.

## Notes

### Competing Interest Statement

The authors have declared no competing interest.

